# Real-World Matched Analysis (N=40 per group) Shows Significantly Improved Healing with Intact Fish Skin Graft vs Standard of Care in Stage 3–4 Pressure Ulcers

**DOI:** 10.64898/2026.04.08.26350429

**Authors:** Hongyu Miao, Ben LeBoutillier, John C. Lantis, Caroline Fife

## Abstract

**Objective:** To evaluate the real-world effectiveness of Intact Fish Skin Graft (IFSG) compared with standard of care (SOC) in the treatment of Stage 3-4 pressure ulcers, using clinically meaningful outcomes including wound healing rate and percent area reduction (PAR).

**Materials and Methods:** A retrospective matched cohort study was conducted using deidentified electronic health record (EHR) data from the U.S. Wound Registry. Patients with Stage 3-4 pressure ulcers treated with IFSG (n=40) were compared to a matched SOC control group (n=40). 1:1 covariate matching was performed to reduce confounding across key patient and wound characteristics, including age, mobility status, comorbidities (e.g., diabetes, peripheral artery disease), and wound features (age, size, location, and depth). Outcomes included healed status, healed or improved rate, and percent area reduction (PAR).

**Results:** The study population represented a high-risk, real-world cohort (n=40 per group), with only 37.5% ambulatory patients and a high prevalence of multiple concurrent wounds. IFSG treatment demonstrated superior clinical outcomes compared to SOC:

- Healed or improved: 67.5% (IFSG) vs 55.0% (SOC) (p=0.0379)
- Healed: 45.5% (IFSG) vs 33.3% (SOC)
- Percent area reduction (PAR): 49% (IFSG) vs 34% (SOC) (p=0.0028)

These findings indicate statistically significant improvements in percent area reduction and in the proportion of wounds that were healed or improved with IFSG. The proportion achieving complete healing was numerically higher with IFSG than with SOC, but this difference did not reach statistical significance.

**Conclusion:** In this real-world matched cohort analysis, Intact Fish Skin Graft demonstrated superior effectiveness compared to standard of care in the management of Stage 3–4 pressure ulcers, with improvements in healing-related outcomes and percent area reduction. These results support the use of IFSG as an effective advanced therapy for hard-to-heal pressure ulcers.

## Introduction

In the United States, pressure ulcers are a major category of chronic, hard-to-heal wound, prevalent in both the acute and long-term care setting. According to data from >90,000 patients in the Pressure Ulcer Prevalence Survey, approximately 12.3% of patients in acute care had pressure ulcers in 2009.^1^ Analysis of the National Minimum Data Set 3.0 showed that of 2.9 million nursing home residents, 8.4% had a stage 2,3,4 or unstageable pressure ulcer.^2^ These percentages translate to millions of pressure ulcers in the U.S. each year and some estimates place the number at 2.5 million or more.^3,4^

It has been estimated that the U.S. healthcare system spends $26.8 billion each year on pressure ulcers acquired in acute care alone.^3^ Action is needed to reduce both the prevalence of pressure ulcers and their associated costs. Besides increased implementation of preventative measures, improved treatments for these chronic wounds are needed to speed and generally increase wound closure.

Biologic tissue grafts have shown positive clinical results when used to treat hard-to-heal chronic wounds, including pressure ulcers. Recently, an interprofessional team of surgeons and nurses specializing in pressure ulcer care has advised using a bioscaffold to support healing when surgical closure was not an option in severe pressure ulcers.^5^ The biologic tissue graft intact fish skin graft (IFSG) has been shown to improve healing in a variety of chronic wounds.^6–8^ IFSG is North Atlantic cod skin that has been minimally processed to remove cells, preserving the extracellular matrix.^9^ This process retains the skin’s natural three-dimensional architecture, mechanical strength, and bioactive constituents, including collagen, proteoglycans, glycoproteins, elastin, lipids, and omega-3 polyunsaturated fatty acids (PUFAs)^10,11^—many of which are essential for effective wound healing in human skin.^12^ The graft’s structure and composition closely mirror those of human skin, providing a structurally and biologically optimized foundation that supports tissue regeneration.^13^ This study seeks to determine whether or not IFSG is effective in improving the healing outcomes of severe pressure ulcers.

Prospective randomized controlled trials (RCTs) have reported Stage 3 and 4 ulcer healing rates of between 29% and 79% within 12 weeks.^14–16^ To accommodate the typical 12–16 week duration of RCTs, subjects with significant comorbid diseases are generally excluded. The healing rates reported from these non-generalizable studies of clinical efficacy are much higher than those observed in the real world.^17^ Indeed, analyses of real-world data reflective of the true patient population have revealed much lower healing rates; for example one real-world study found healing rates of only 31.3% and 12.9% over 6 months for Stage 3 and Stage 4 ulcers, respectively,^18^ and another real-world study found that over the course of 12 months, just 41% of Stage 3 ulcers and 21% of Stage 4 ulcers healed.^19^ Thus, when evaluating whether a particular treatment improves the outcome of pressure ulcers in clinical practice, it is vital to analyze effectiveness from real-world data.

Here we probe the effectiveness of IFSG using real-world EHR data derived directly from point-of-care clinical documentation. We compare the healing rate of Stage 3 and 4 pressure ulcers in patients treated with IFSG versus a matched cohort of patients treated without IFSG. The IFSG-treated and control groups are matched on 15 patient and wound characteristics known to influence the odds of healing.

## Materials and Methods

### Source of Real-World Data

Real-world data were sourced from the U.S. Wound Registry (USWR), which is comprised of deidentified patient data transferred electronically from a wound-care specific, highly structured electronic health record (Intellicure, LLC, The Woodlands, TX). Over 4,000 practitioners across 34 U.S. States and Puerto Rico utilize the Intellicure EHR. Although the data are proprietary, they may be made available through data-use agreements, as described in the Data Availability Statement. To extract clinical data specific to pressure ulcers from USWR, the ICD10 codes were used to identify pressure ulcers between the years 2018 (Jan. 1^st^) and 2025 (Jun. 30^th^).

### Identifying the IFSG-Treated Group for Analysis

First, all IFSG-treated pressure ulcers were extracted from the USWR. Minimal inclusion/exclusion criteria were applied to improve internal validity and ensure clinically meaningful comparisons. In pressure ulcers treated with one or more other skin substitutes in addition to IFSG, only ulcers in which IFSG was the final skin substitute used were included, allowing outcomes to be more directly attributed to IFSG and reduce potential treatment contamination. Additionally, pressure ulcers in unusual locations were excluded due to the impossibility of identifying an adequate cohort match (e.g., “breast” and “vagina”) Thirdly, wounds ≤ 1 cm^2^ were excluded because, in very small wounds, even minimal absolute reductions in area can translate into disproportionately large percentage area reductions, potentially influencing estimates of treatment effect. Also, wounds ≤ 1 cm^2^ are difficult to measure accurately. Finally, when multiple pressure ulcers were present on a given patient, we analyzed an index ulcer defined as the largest pressure ulcer to keep the wound-patient ratio 1:1. Because we wanted to study the effectiveness of IFSG on the severe pressure ulcers, we included only full-thickness pressure ulcers corresponding to Stages 3 and 4. The final IFSG-treated group consisted of 40 pressure ulcers on 40 patients.

### Identifying the Control Group for Analysis

Pressure ulcers that were never treated with any skin substitute comprised the standard of care (SOC) control cohort. IFSG-treated pressure ulcers were matched 1:1 with SOC ulcers based on 15 covariates for a total SOC control cohort of 40 Stage 3 and 4 pressure ulcers on 40 patients. The covariate matching is discussed in more detail in the below section entitled, “Matching Model.”

### Matching Model

To minimize confounding, covariates were matched between the IFSG-treated group and control group using Mahalanobis distance and nearest neighbor with the tool R v4.5.1, MatchIt package v4.7.2. For matched continuous covariates, the signed rank test was used to compare between IFSG-treated and control groups. The McNemar test was used to compare matched categorical covariates between groups.

IFSG-treated and control groups were matched 1:1 on 15 patient and wound covariates. Matched patient covariates include patient age, sex, method of arrival to appointments, number of concomitant wounds, and whether the patient has each of the following comorbidities: diabetes, chronic kidney disease (CKD), congestive heart failure (CHF), peripheral arterial disease (PAD), autoimmune disease, dementia, and paralysis. Matched wound covariates include age, size, location on the body, and deepest tissue type exposed.

These matched variables have been shown to affect the odds of healing specifically in pressure ulcers (patient age, method of arrival to appointments, number of concomitant wounds, diabetes, CKD, PAD, autoimmune disease, dementia, paralysis, wound age, wound size, wound location on the body, and deepest tissue type exposed) or in chronic wounds in general (patient sex and CHF).

Note that the patient age was adjusted to the date of the first visit for a particular wound. The wound location was determined based on ICD-10 codes (L89.0-L89.9). The wound age was calculated from the (self-reported) onset time to the first application of IFSG (or until the index date for the control group).

### Determination of Index Date

In the IFSG-treated group, the index date was set as the day of first application with IFSG for each wound, defined as Day 0 and from which all times to a specific outcome were calculated. For the control group, the index date was set to the day at which the control pressure ulcer was closest in area to its matched IFSG-treated ulcer.

### Healing Outcome Imputed from Percentage Area Reduction

If the percentage of wound size reduction was greater than or equal to 97% at the last measurement, the outcome was imputed as “Healed.” If, over a period of 100 days (since real data suggest that the average wound time in service is approximately this long), the percentage of wound size change was between ±10%, the outcome was imputed as “No Change.” If over 100 days, the percentage wound size change was less than -10%, the outcome was imputed as “Healing” and if over this timeframe the percentage wound size change was greater than +10%, the outcome was imputed as “Worse.” Serial wound area measurements documented in the EHR between the index date and the last recorded assessment were used to compute PAR and assign these categories; when intermediate measurements were missing, the last observed area was carried forward (LOCF) until the next documented assessment or the end of follow-up. The time frame for determining wound PAR and healing spanned from the index date to the last recorded wound assessment and measurement.

## Results

The matched patient covariates were paired either perfectly or almost perfectly; those paired almost perfectly were not statistically different between IFSG-treated and control groups (Table 1). As shown in Table 1, the mean ages in the IFSG-treated group and control group were 66.60 and 65.62, respectively. The proportion of patients ≥65 was similar and did not differ statistically between the two groups. Patients skewed male at 62.5%. Several matched comorbidities had an incidence of one quarter or higher; 27.5% of patients had diabetes, 25.0% of patients had PAD, and 25.0% of patients had autoimmune disease. Additionally, 12.5% of patients had chronic kidney disease, 10.0% of patients had congestive heart failure, and 7.5% had dementia. 35.0% and 32.5% of patients (in the IFSG-treated group and control group, respectively) were paralyzed. The method of arrival to appointments, a surrogate for overall mobility status, showed that more than half of patients used a wheelchair, 5.0% used a stretcher or were bedridden, and only 37.5% were ambulatory. The vast majority of patients had other wounds in addition to the index wound, with 72.5% and 80% of the IFSG-treated group and control group, respectively, having one or more other wounds present at the same time as the index wound (Figure 1).

**Table 1:** Matched Patient Covariates, Note that the slight differences between the cohorts in age and prevalence of paralysis were not statistically significant.

| **Matched Covariate** | **IFSG-treated group** | **Control group** |
| --- | --- | --- |
| **Age** [Mean (SD)] | 66.60 (18.03) | 65.62 (17.69) |
| **Male** | 25 (62.5%) | 25 (62.5%) |
| **Diabetes** | 11 (27.5%) | 11 (27.5%) |
| **Chronic Kidney Disease** | 5 (12.5%) | 6 (12.5%) |
| **Autoimmune Disease** | 10 (25.0%) | 10 (25.0%) |
| **Congestive Heart Failure** | 4 (10.0%) | 4 (10.0%) |
| **Dementia** | 3 (7.5%) | 3 (7.5%) |
| **Paralysis** | 14 (35.0%) | 13 (32.5%) |
| **Peripheral Artery Disease** | 10 (25.0%) | 10 (25.0%) |
| **Arrival Method** |  |  |
| Ambulatory | 15 (37.5%) | 15 (37.5%) |
| Stretcher/Bedridden | 2 (5.0%) | 2 (5.0%) |
| Wheel chair | 23 (57.5%) | 23 (57.5%) |
| **Number of Concomitant Wounds** | See Figure 1 | See Figure 1 |

**Figure 1:**
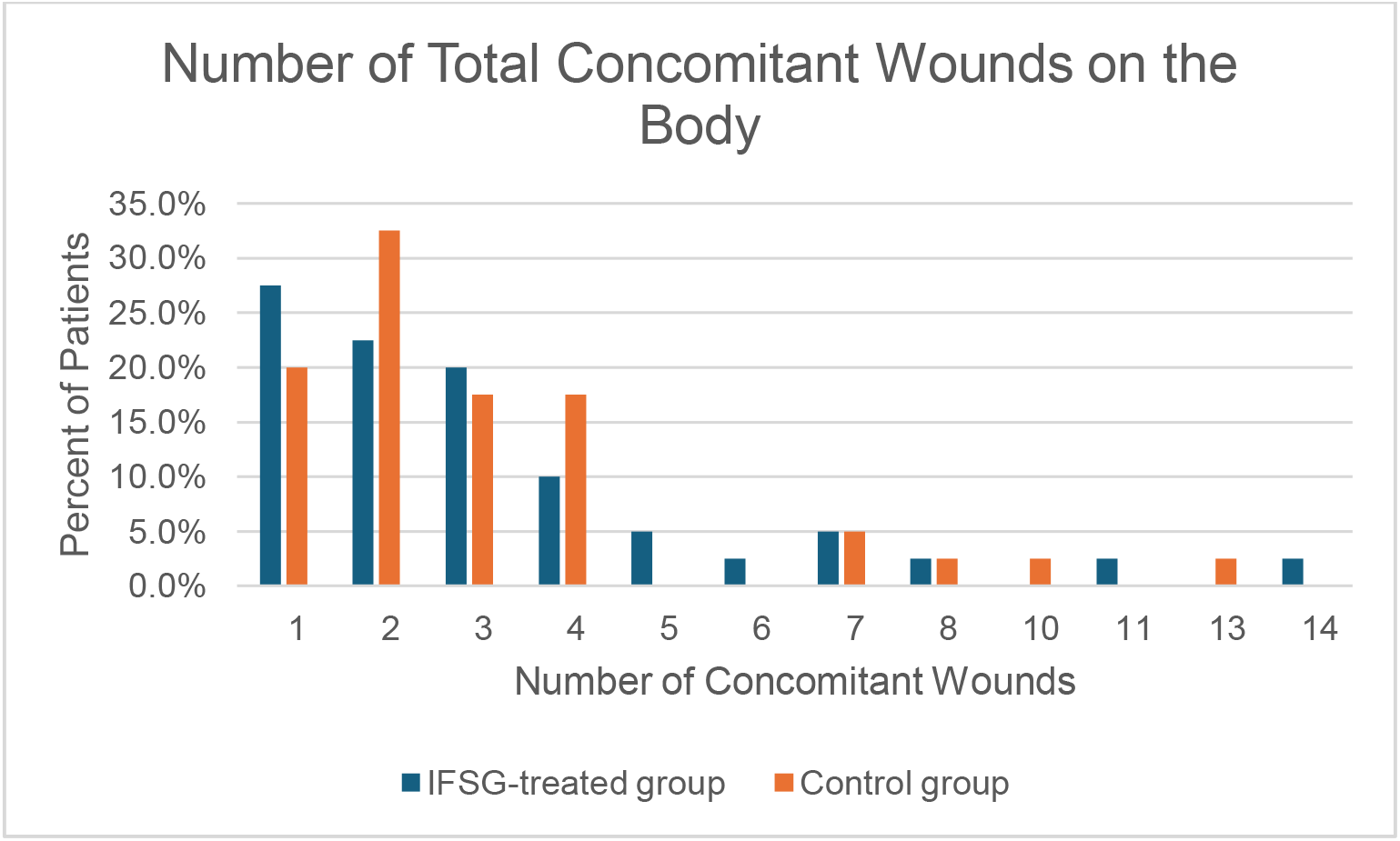
The total number of concomitant wounds was well matched between the IFSG-treated and control groups.

Unmatched patient characteristics were also well balanced and were not statistically different between groups (Table 2). Patients were predominantly White (90.0% and 96.6%) with some Black or African American and one patient who identified as “Other,” although data on race was not available for all patients. The average BMI for both groups was in the “Overweight” category and 45.0% and 30.0% of patients (in the IFSG-treated group and the control group, respectively) were in the “Obese” category. Unmatched comorbidities can be seen in Table 2. Note that 27.5% of IFSG-treated patients had a history of osteomyelitis, while only 7.5% of control patients did, although this difference between groups was just below the level of statistical significance (p=0.0614).

**Table 2:** Additional Patient Demographics, Note that the differences between the cohorts were not statistically significant.

| **Other Patient Characteristics** | **IFSG-treated group** | **Control group** |
| --- | --- | --- |
| **Race** |  |  |
| Black or African American | 2 (6.7%) | 1 (3.4%) |
| Other | 1 (3.3%) | 0 (0.0%) |
| White | 27 (90.0%) | 28 (96.6%) |
| Missing | 10 | 11 |
| **BMI** |  |  |
| Mean (SD) | 27.22 (6.43) | 26.94 (8.93) |
| Median (Q1, Q3) | 26.70 (23.93, 30.43) | 26.20 (20.12, 32.02) |
| Missing | 2 | 2 |
| **Obesity** | 18 (45.0%) | 12 (30.0%) |
| **Tobacco Use** | 5 (12.5%) | 5 (12.5%) |
| **Depression** | 1 (2.5%) | 4 (10.0%) |
| **Atrial Fibrillation** | 6 (15.0%) | 4 (10.0%) |
| **Osteomyelitis** | 11 (27.5%) | 3 (7.5%) |
| **Hyperlipidemia** | 9 (22.5%) | 7 (17.5%) |
| **Hypertension** | 11 (27.5%) | 16 (40.0%) |

The matched wound covariates are shown in Table 3. All wounds were either Stage 3 or Stage 4 pressure ulcers, with 75.0% Stage 3 and 25.0% Stage 4. In the Stage 4 wounds, the deepest tissues exposed included muscle (12.5%), tendon (7.5%), and bone (5.0%). Wounds were at a range of anatomical locations (Figure 2), though those occurring on the heel or midfoot made up the largest group (30.0%). Wound age, calculated as time from the onset date to the index date, was matched as closely as possible, but still ended up being statistically different between the two groups (p= 0.0081). On average, the IFSG-treated wounds were older than the control wounds; mean wound age was 360.00 days for IFSG-treated wounds and 233.62 days for control wounds. Baseline wound size was closely matched between the two groups. On the index date, the mean wound area was 8.99 cm^2^ for the IFSG-treated group and 9.03 cm^2^ for the control. Although not a covariate used for matching, the amount of granulation present on the index date was made to be exceedingly similar between groups (Figure 3).

**Table 3.** Matched Wound Covariates, * The difference in wound age at baseline between the two cohorts reached statistical significance (p=0.0081).

| **Matched Covariate** | **IFSG-treated group** | **Control group** |
| --- | --- | --- |
| **Deepest Tissue Exposed at Baseline** |  |  |
| BONE | 2 (5.0%) | 2 (5.0%) |
| MUSCLE | 5 (12.5%) | 5 (12.5%) |
| SUBCUTANEOUS | 30 (75.0%) | 30 (75.0%) |
| TENDON | 3 (7.5%) | 3 (7.5%) |
| **Wound Age at Baseline \*** |  |  |
| Mean (SD) | 360.00 (445.69) | 233.62 (347.59) |
| Median (Q1, Q3) | 197.50 (89.75, 473.50) | 91.50 (49.50, 295.50) |
| **Wound Size at Baseline** |  |  |
| Mean (SD) | 8.99 (11.57) | 9.03 (12.81) |
| Median (Q1, Q3) | 4.70 (1.65, 10.12) | 4.34 (1.55, 13.43) |
| **Wound Stage** |  |  |
| Stage 3 | 30 (75.0%) | 30 (75.0%) |
| Stage 4 | 10 (25.0%) | 10 (25.0%) |
| **Wound Location** | See Figure 2 | See Figure 2 |

**Figure 2:**
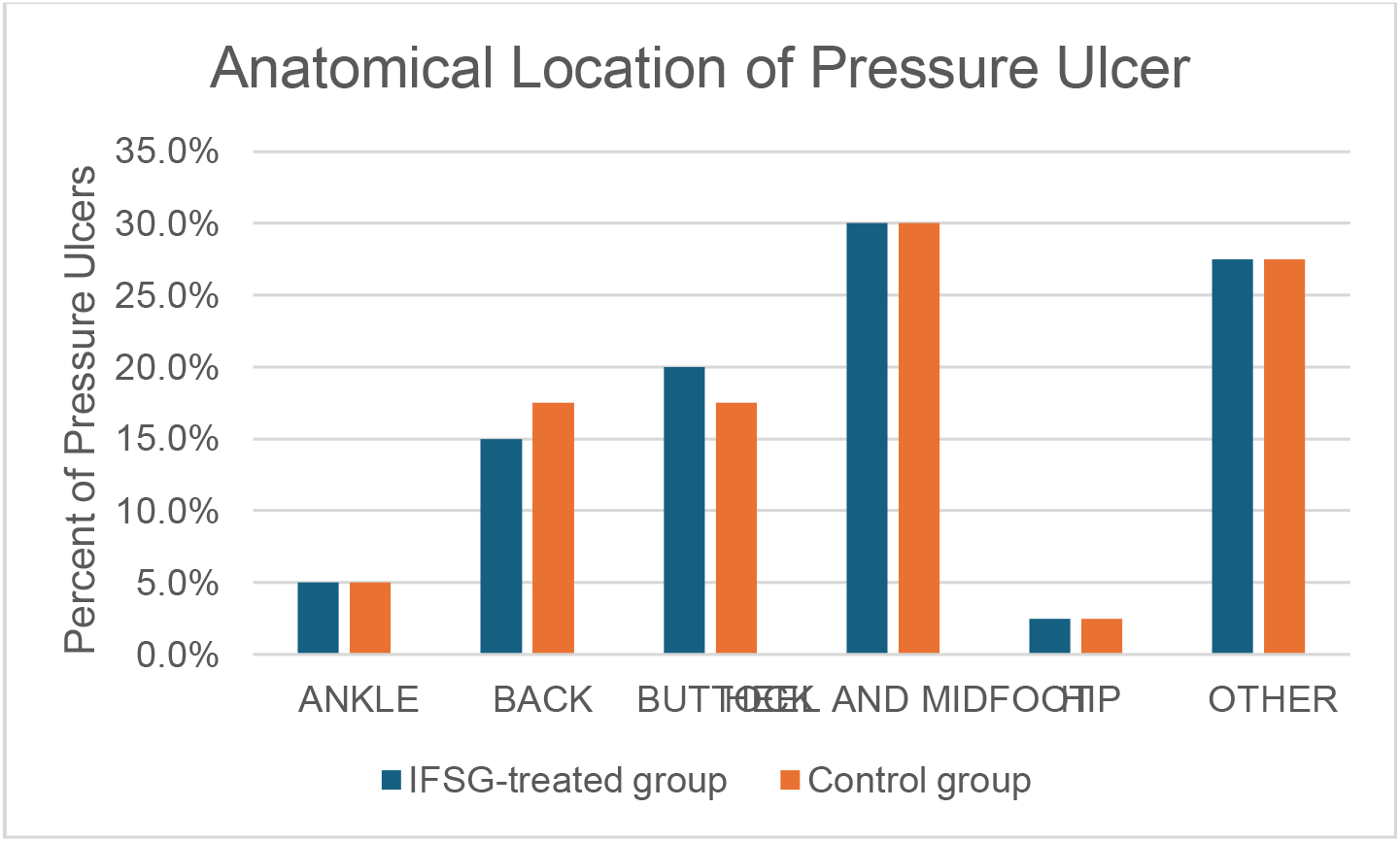
Pressure ulcer location was well matched between the IFSG-treated and control groups.

**Figure 3:**
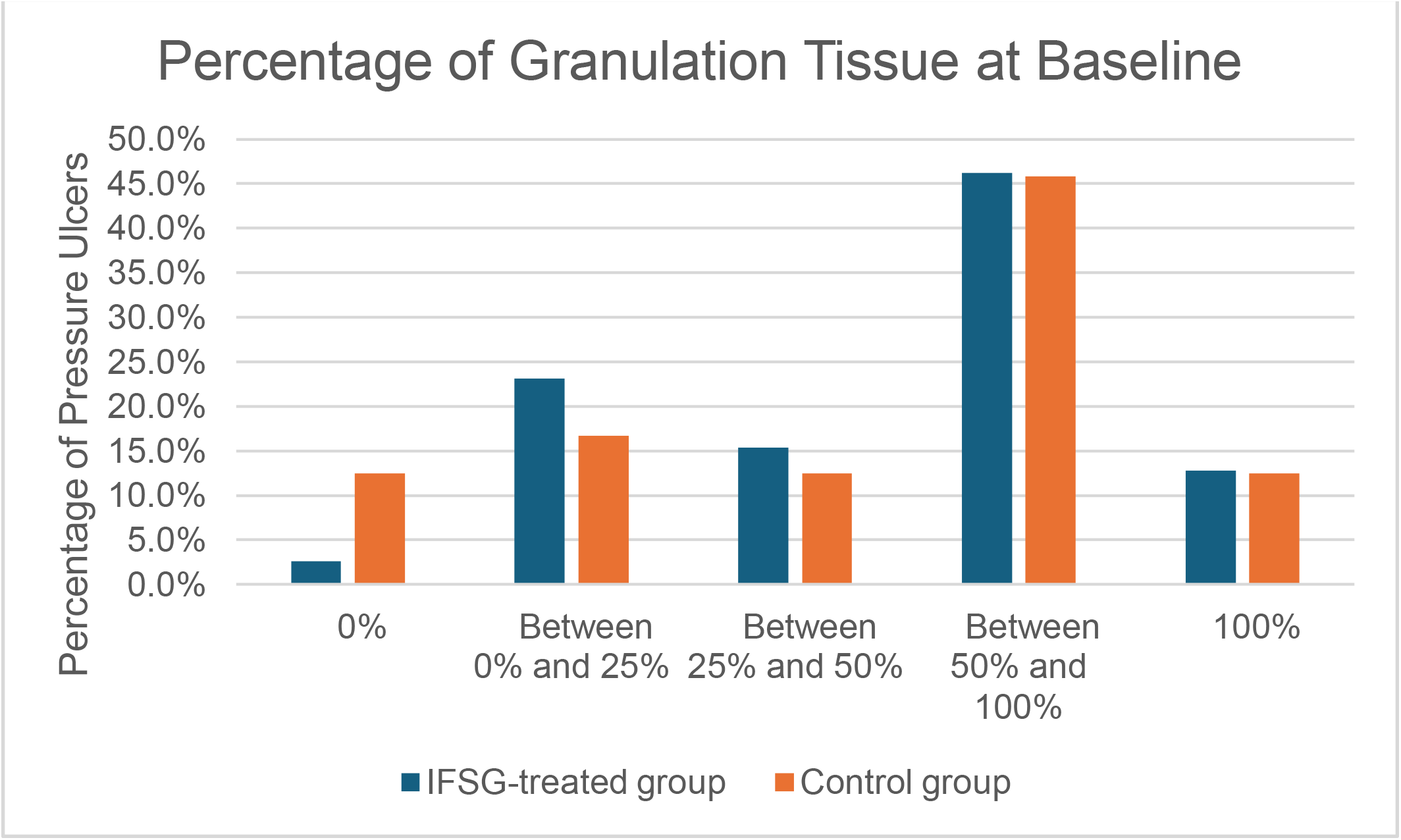
The IFSG-treated and control groups had similar baseline degrees of wound granulation.

The average time-in-service before the first application of IFSG was 152.10 days and the average time-in-service before the index date for the control was 80.25 days, a difference of 71.85 days that was statistically significant (paired t-test, p=0.0015).

The total proportion healed for the IFSG-treated group was 45.5%, while the total proportion healed for the control group was 33.3%, a difference of 11.7% (Table 5). This absolute gain with IFSG did not reach statistical significance in the paired analysis, which is expected given the modest number of matched pairs (n=40 per arm) and the resulting limited power to detect differences in binary complete healing, despite the numerical improvement. When both healed wounds and wounds that improved are included together, the IFSG-treated group had 67.5% of wounds heal or improve while the control group had 55.0% of wounds heal or improve, a difference of 12.5%, which is statistically significant (paired t-test, p=0.0379) (Table 6). On average, ulcers treated with IFSG reduced in size by 49% and ulcers in the control group reduced in size by 34% (Table 4). This 15% difference is statistically significant (paired t-test, p=0.0028).

**Table 4.** Percentage Area Reduction, *The difference in PAR was statistically significant (paired t-test, p= 0.0028)

| **Descriptive Statistic** | **IFSG-treated group** | **Control group** |
| --- | --- | --- |
| Mean (SD)* | -49% (152%) | -34% (48%) |
| Median (Q1, Q3) | -0.99 (-1.00, -0.59) | -0.24 (-0.80, 0.00) |

**Table 5.**
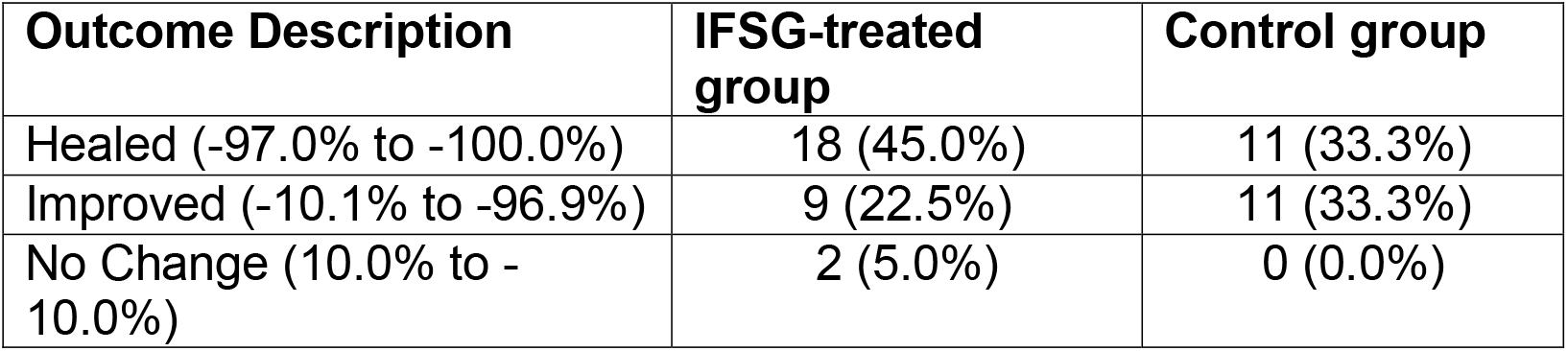
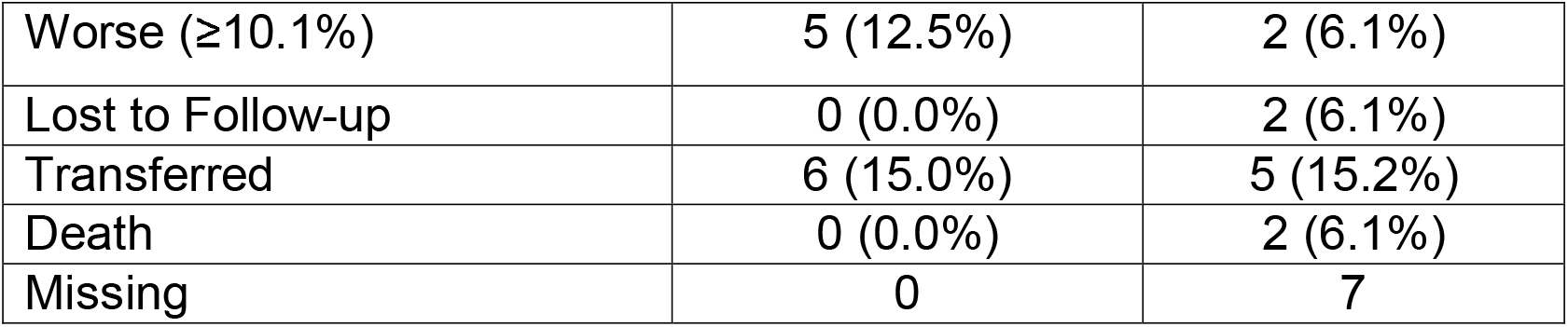
Percentages are PARs with negative values corresponding to a shrinking wound and positive values indicating wound expansion.

**Table 6.** *The difference in the rate of Healed or Improved was statistically significant (paired t-test, p= 0.0379)

| Outcome Type | IFSG-treated group | Control group |
| --- | --- | --- |
| **Healed or Improved\***<br>(-10.1% to -100.0%) | 27 (67.5%) | 22 (55.0%) |
| **No Change or Worse**<br>(-10.0% to 10.0% or >10.0%) | 7 (17.5%) | 2 (5.0%) |
| **Unknown (transfer, lost to follow-up, death, or missing)** | 6 (15.0%) | 16 (40.0%) |

## Discussion

The results of this study demonstrate that treatment with IFSG provides statistically significant advantages over standard of care (SOC) alone for Stage 3 and 4 pressure ulcers with respect to percent area reduction (PAR) and the proportion of wounds that were healed or improved. The proportion achieving complete healing was higher numerically with IFSG (45.5%) than with SOC (33.3%), but this difference was not statistically significant, consistent with limited power in a matched sample of 40 wound pairs even when an absolute gain of this magnitude is clinically meaningful. The control group achieved a healing rate of 33.3% over an average of 16 weeks—a figure consistent with real-world longitudinal data showing six-month healing rates between 12.9% and 31.3%.^18^ In contrast, the IFSG-treated group showed a 45.5% healing rate. When expanding the clinical success metric to include wounds that were “Healed or Improved,” the IFSG group reached 67.5% compared to 55.0% in the control group, a 12.5% difference that was statistically significant (p=0.0379). Further, when percent area reduction (PAR) is analyzed, IFSG treatment also shows superiority to SOC; wounds treated with IFSG saw an average size reduction of 49%, whereas the control group reduced in size by an average of 34%. This 15% difference is statistically significant (paired t-test, p=0.0028).

Our data show that clinicians in this real-world setting waited an average of 152.1 days (over 21 weeks) before applying IFSG to pressure ulcers, suggesting that conservative measures were attempted prior to initiating advanced therapy. The observed improvement despite this substantial delay supports the effectiveness of IFSG; however, one might reasonably question whether earlier application could have produced even better outcomes.

The ICD10-CM provides pressure ulcer codes specific to ulcer stage and location for the only the following specific anatomical areas: head, elbow, heel, ankle, back, upper back, sacrum, hip, buttock, and contiguous site of back, buttock and hip. However, these are not the only anatomical locations where pressure ulcers are documented to occur in the real world. USWR data show that 20.9% of all pressure ulcers in the real world are in anatomical locations for which there is no specific ICD10 code, based on free-text analysis of clinical data, and of this group, 93.3% are documented to occur on the foot, toes and leg. Other documented locations include the abdomen, genitalia, face, and upper extremity in areas other than the elbow. Thus, the prevalence of “other” locations in IFSG treated pressure ulcers is consistent with USWR anatomical location data. In this study, 30% of IFSG-treated pressure ulcers were located on the heel or midfoot. Based on analysis of more than 140,000 pressure ulcers on the USWR, approximately 20.3% of pressure ulcers are on the heel.

As is expected in a real-world patient population, patients examined in this study were characterized by significant yet common comorbidities known to make healing more challenging, including peripheral artery disease (25.0%), autoimmune disease (25.0%), and diabetes (27.5%).^20,21^ Furthermore, only 37.5% of patients were ambulatory, with the majority being wheelchair or bedbound—a known negative prognostic factor for chronic wound resolution.^20,21^ A total of 72.5% IFSG-treated patients and 80.0% control patients had more wounds than the index pressure ulcer present simultaneously. The presence of multiple wounds simultaneously is a common occurrence in chronic wound patient populations and is negatively correlated with healing.^21^

Confounding can be a challenge in studies using real-world data. Matching variables known or predicted to affect outcomes between groups is one method of minimizing confounding and increasing study robustness. Here we matched 15 patient and wound covariates known to negatively affect healing between IFSG-treated and control groups, a degree of matching not previously accomplished in real world pressure ulcer data. Additionally, other patient and wound characteristics were well-balanced between the two groups. Notably, the few patient and wound variables that did differ between groups tended to bias against the IFSG-treated group. For instance, the IFSG-treated group had significantly older wounds at baseline (360 days vs. 233.6 days, p=0.0081) and 27.5% of IFSG-treated patients had a history of osteomyelitis while only 7.5% of control patients did, a trend that bordered on statistical significance (p= 0.0614). The fact that IFSG-treated wounds had greater healing and PAR despite these potential biases further supports the effectiveness of IFSG.

## Conclusion

The findings of this study demonstrate that IFSG is a highly effective advanced therapy for the management of Stage 3 and 4 pressure ulcers in a real-world clinical setting. IFSG was associated with statistically significant increases in both the proportion of wounds healed or improved (67.5% vs 55.0%) and PAR (49% vs 34%). Complete healing was more common numerically with IFSG than SOC (45.5% vs 33.3%) but did not differ significantly at conventional thresholds, likely reflecting limited statistical power with 40 matched pairs rather than absence of clinical benefit. Larger real-world or randomized evaluations, including subgroup analyses by ulcer stage and patient risk profile, are warranted to confirm and extend these findings.

## Data Availability

Data produced in the present study may be available upon reasonable request to the authors.

